# Deep-Learning Based Contrast Boosting Improves Lesion Visualization and Image Quality: A Multi-Center Multi-Reader Study on Clinical Performance with Standard Contrast Enhanced MRI of Brain Tumors

**DOI:** 10.1101/2025.06.12.25329347

**Authors:** Srivathsa Pasumarthi Venkata, Thomas Campbell Arnold, Sonia Colombo Serra, Jeffrey. D. Rudie, Jalal. B. Andre, Ronny Elor, Praveen Gulaka, Ajit Sankaranarayanan, Gunter Erb, Greg Zaharchuk

**Author notes:** Corresponding author: Srivathsa Pasumarthi Venkata.

## Abstract

**Background:** Gadolinium-based Contrast Agents (GBCAs) are used in brain MRI exams to improve the visualization of pathology and improve the delineation of lesions. Higher doses of GBCAs can improve lesion sensitivity but involve substantial deviation from standard-of-care procedures and may have safety implications, particularly in the light of recent findings on gadolinium retention and deposition.

**Purpose:** To evaluate the clinical performance of an FDA cleared deep-learning (DL) based contrast boosting algorithm in routine clinical brain MRI exams.

**Methods:** A multi-center retrospective database of contrast-enhanced brain MRI images (obtained from April 2017 to December 2023) was used to evaluate a DL-based contrast boosting algorithm. Pre-contrast and standard post-contrast (SC) images were processed with the algorithm to obtain contrast boosted (CB) images. Quantitative performance of CB images in comparison to SC images was compared using contrast-to-noise ratio (CNR), lesion-to-brain ratio (LBR) and contrast enhancement percentage (CEP). Three board-certified radiologists reviewed CB and SC images side-by-side for qualitative evaluation and rated them on a 4-point Likert scale for lesion contrast enhancement, border delineation, internal morphology, overall image quality, presence of artefacts, and changes in vessel conspicuity. The presence, cause, and severity of any false lesions was recorded. CB results were compared to SC using Wilcoxon signed rank test for statistical significance.

**Results:** Brain MRI images from 110 patients (47 ± 22 years; 52 Females, 47 Males, 11 N/A) were evaluated. CB images had superior quantitative performance than SC images in terms of CNR (+634%), LBR (+70%) and CEP (+150%). In the qualitative assessment CB images showed better lesion visualization (3.73 vs 3.16) and had better image quality (3.55 vs 3.07). Readers were able to rule out all false lesions on CB by using SC for comparison.

**Conclusions:** Deep learning based contrast boosting improves lesion visualization and image quality without increasing contrast dosage.

**Key Results:**

- In a retrospective study of 110 patients, deep-learning based contrast boosted (CB) images showed better lesion visualization than standard post-contrast (SC) brain MRI images (3.73 vs 3.16; mean reader scores [4-point Likert scale])
- CB images had better overall image quality than SC images (3.55 vs 3.07)
- Contrast-to-noise ratio, Lesion-to-brain Ratio and Contrast Enhancement Percentage for CB images were significantly higher than SC images (+729%, +88% and +165%; p < 0.001)

**Summary Statement:** Deep-learning based contrast boosting achieves better lesion visualization and overall image quality and provides more contrast information, without increasing the contrast dosage in contrast-enhanced brain MR protocols.

## Introduction

Gadolinium-based Contrast Agent (GBCA) enhanced brain MRI is used to detect a wide range of pathological processes[1–4], which would be otherwise undetectable through other methods[5,6], and it is the primary method for diagnosis, treatment and surgical planning for brain tumors[7, 8]. The administered contrast dose is proportional to the T1 signal intensity[9] and studies have shown that higher dosage yields better sensitivity, especially in the detection of smaller lesions[10]. However, recent findings on long-term gadolinium retention/deposition in tissues[11] and gadolinium water contamination[12] have raised safety and environmental concerns over increasing the contrast dosage, even though no clinical signs could be assigned to these findings [13].

Multiple approaches have been researched to improve the diagnostic accuracy and sensitivity of contrast enhanced (CE) MRI without increasing the dose. Higher relaxivity GBCAs have been recently introduced[14–16] which are being used at lower dose levels to achieve comparable lesion visualization. Non-contrast enhanced MRI protocols such as arterial spin labeling (ASL)[17] and time-of-flight MR angiography (TOF-MRA)[18] have also been considered as alternatives. Numerous studies have found deep learning (DL) algorithms can be applied to reduce contrast dosage while maintaining standard-of-care contrast enhancement[19–23]. More recently, researchers have proposed repurposing dose reduction algorithms to maximize the contrast enhancement present in standard dose CE MRI[24,25].

This present study proposes a multi-reader multi-center clinical performance evaluation of a DL algorithm that boosts contrast information present in standard CE brain MRI. We first trained a DL model to predict standard-contrast (SC) enhanced images from pre-contrast and low-dose (10% of standard dose) image inputs. We then applied this model to pre-contrast and SC images to obtain contrast boosted (CB) images. Lesion visualization performance of CB images was quantitatively assessed using metrics and qualitatively evaluated through a multi-reader study. Readers also rated SC and CB images on image quality and the presence of artifacts. Readers also scored images on their extent of vessel conspicuity and its impact on diagnosis. We further analyzed the frequency, cause, and severity of any false lesions introduced by the algorithm. Finally, we quantitatively assess if CB images have an appearance of higher-dose images through a physics-guided dose concentration and CNR analysis.

## Materials and Methods

All patient studies in this retrospective multi-center study were obtained with IRB approval and informed consent. Collection and analysis of demographic, clinical and radiologic data was done with informed written consent.

### Patients

A total of 674 cases (334 Females, 328 Males, 12 N/A; 51 ± 17 years) from six institutions were considered for inclusion in this retrospective study. Images were acquired between April 2017 and Dec 2023. Patients from different institutions underwent clinical brain MRI examination using different scanner manufacturers, vendors and scanning protocols. The clinical indications included a wide range of pathologies requiring GBCA contrast administration. Patients were categorized into 10 clinical indications (listed in Table 1). From this database, 110 patient studies (47 ± 22 years; 52 Females, 47 Males, 11 N/A) were sampled for evaluation, accounting for distribution in terms of patient demographics and scan parameters. Fig. 1 shows the inclusion criteria for this study.

**Fig 1:**
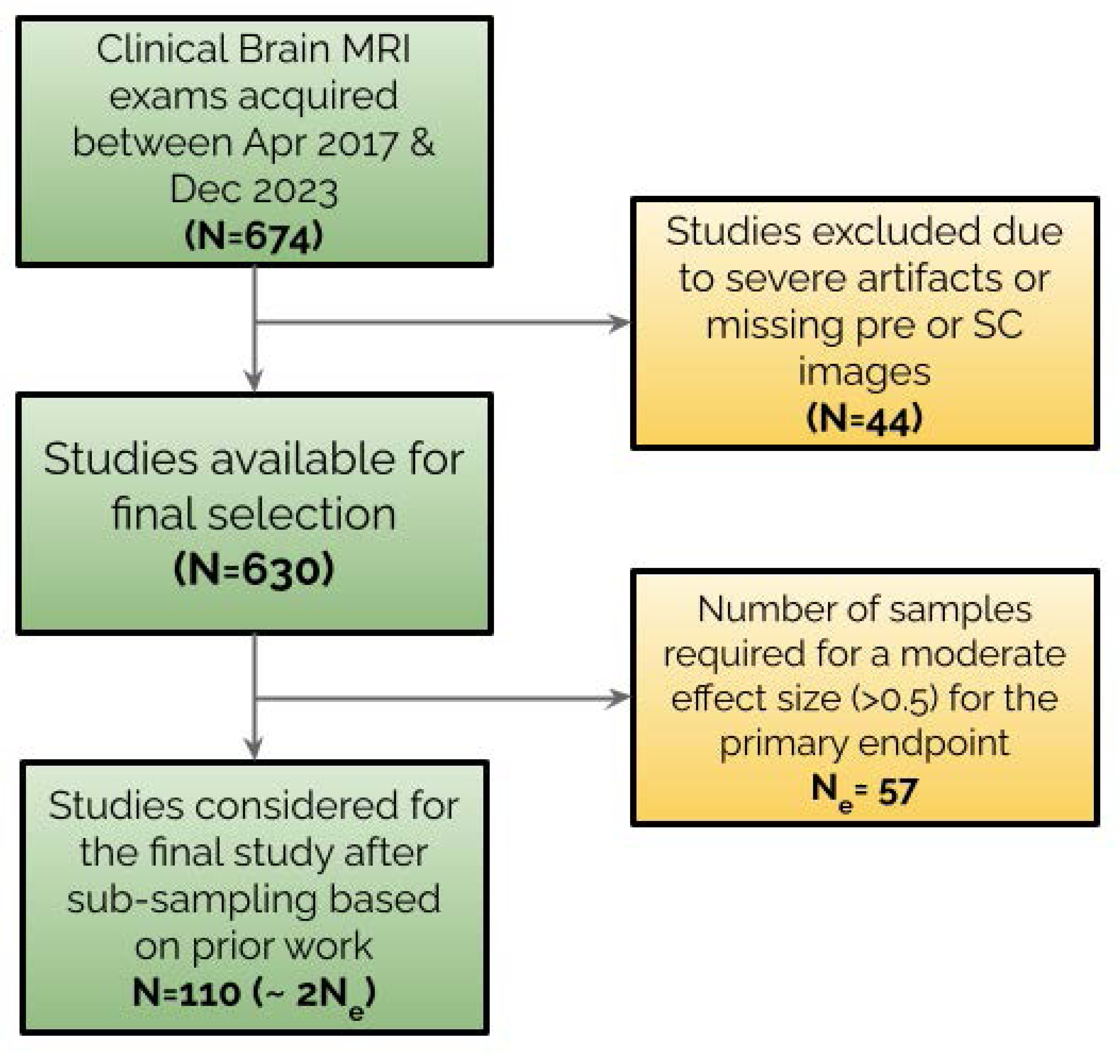
Inclusion criteria and sampling strategy for the evaluation of the contrast-boosting Deep Learning algorithm, based on patient demographics, clinical indications and scanning parameters.

**Table 1:**
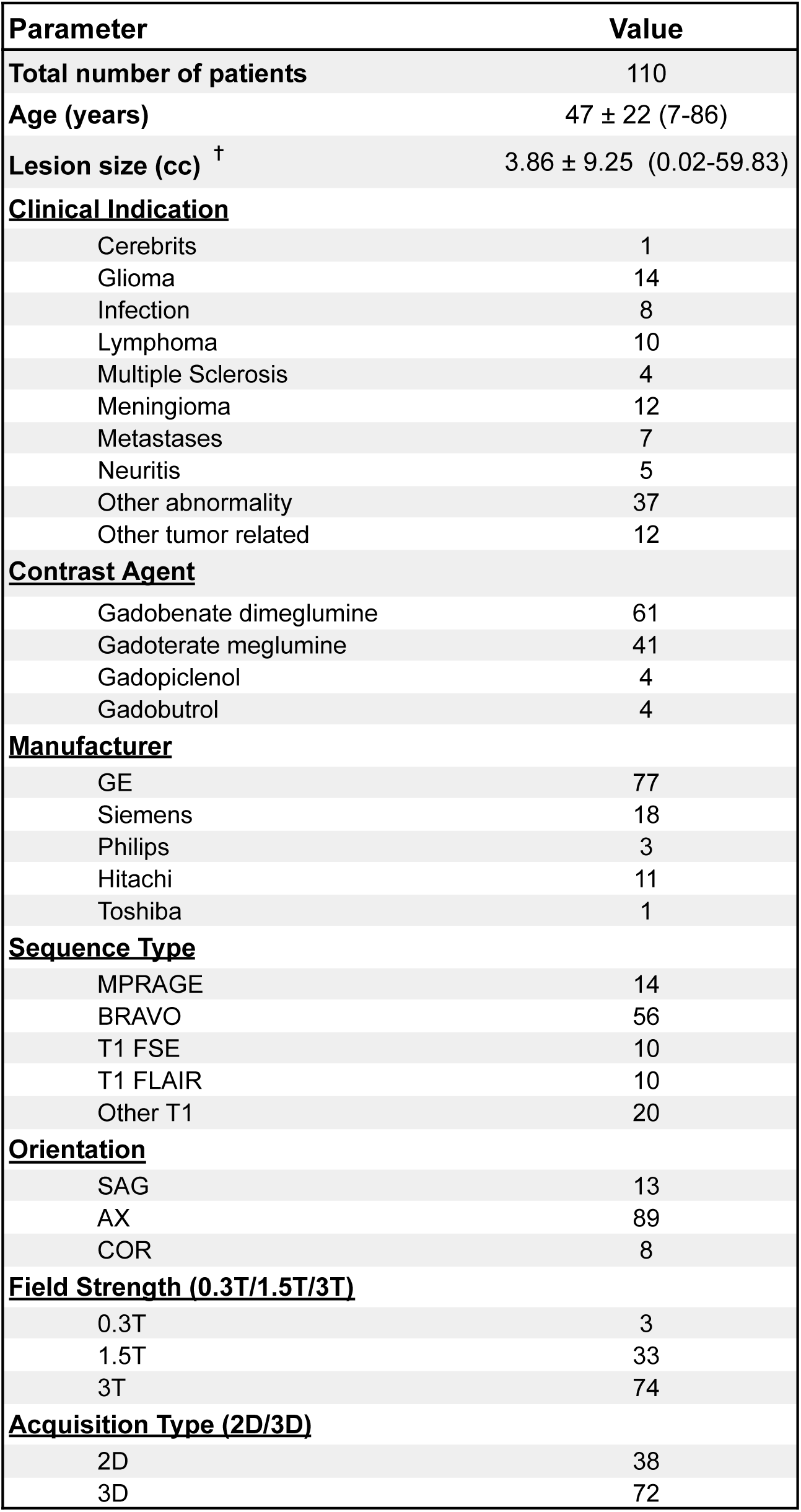
Distribution of exams based on patient sex, clinical indication and different scanning parameters - contrast agent, orientation, manufacturer, field strength, sequence type and 2D vs 3D. ✝ - the protocol for lesion size measurement is described in the Supplementary Material.

The sample size was estimated based on results from a smaller prior study that was conducted. The effect size of the primary qualitative endpoint (lesion contrast enhancement), from our prior study was 0.85. Accounting for enough statistical power to detect a moderate effect size (> 0.5), we needed a minimum sample size of N=57. Thus we chose roughly twice the value and arrived at N=110. Within the sample size, we ensured that subgroups of interest have a minimum representation met to demonstrate generalizability of the algorithm across the patient population and clinical use. Therefore, from the 630 available patient studies, a random sampling was done within each subgroup to arrive at the final evaluation dataset. Table 1 shows the detailed patient data distribution.

### Deep Learning Algorithm

We used pre-contrast and standard post-contrast (SC) images as inputs to generate contrast-boosted (CB) images by applying an FDA-cleared algorithm *(Product A, Company Y and Company Z)*. The algorithm leverages a U-Net based DL network that was previously proposed [20] for contrast dose reduction. The dose reduction model was trained to predict standard dose contrast enhanced brain MRI images from pre-contrast and low-dose (10% of standard dose) images. The dose reduction model performs a non-linear transform of contrast difference between the two input images to achieve contrast boosting. We repurposed the dose reduction model to perform contrast boosting of standard dose images by using pre-contrast and SC images as model inputs. Fig. 2 shows the overall workflow of the contrast boosting algorithm, which includes an FDA-cleared denoising module (*Product B, Company Z*) [26] to input pre-contrast and SC images as shown. The output images from the contrast boost algorithm were also processed with the denoising module to enhance the image quality of the CB images.

**Fig 2:**
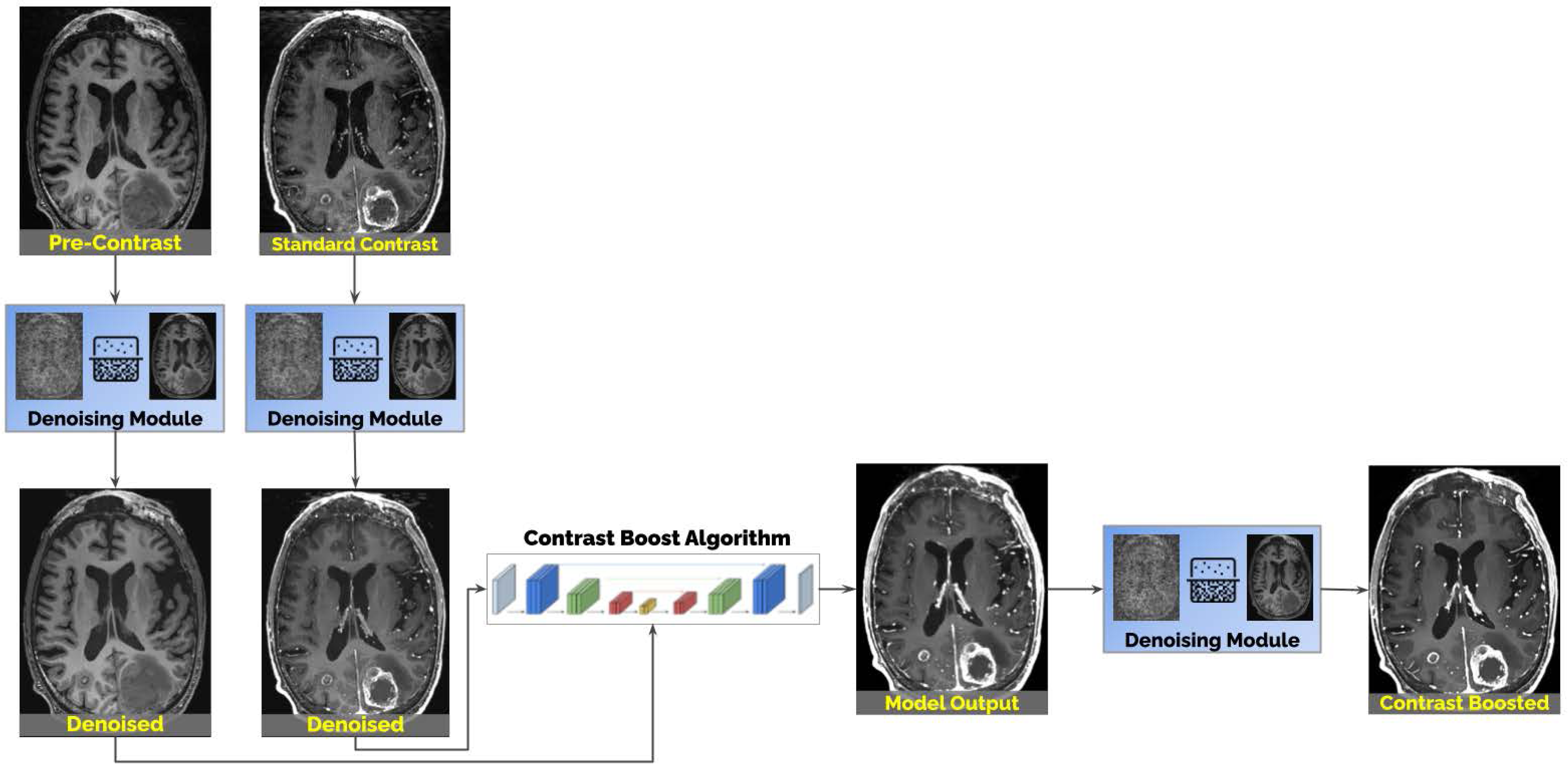
Overall workflow of the deep learning algorithm for contrast boosting. A denoising module is applied on the model inputs and the output.

### Quantitative Assessment

For the quantitative assessment, bounding box ROIs were manually drawn by an independent board-certified radiologist J.S., with 10 years of experience. The quantitative assessment endpoints require two ROIs for each patient study: i) a region of interest around the largest enhancing lesion (𝑅𝑂𝐼_𝑙𝑒𝑠𝑖𝑜𝑛_) and ii) a region of interest of similar dimensions around parenchymal tissue on the same slice as the lesion (𝑅𝑂𝐼_𝑏𝑟𝑎𝑖𝑛_). In the absence of an enhancing lesion, the superior sagittal sinus was used as a surrogate for 𝑅𝑂𝐼_𝑙𝑒𝑠𝑖𝑜𝑛_ to assess contrast enhancement[27]. To quantify the contrast boosting effect of the DL algorithm, we used three quantitative metrics as defined below: i) contrast-to-noise ratio (CNR), ii) lesion-to-brain ratio (LBR), and iii) contrast enhancement percentage (CEP).

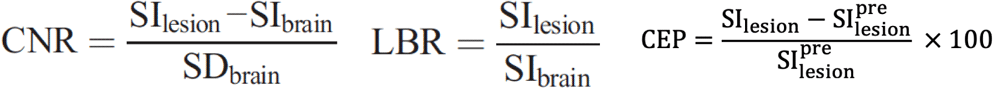

Here, 𝑆𝐼_𝑙𝑒𝑠𝑖𝑜𝑛_ denotes the mean signal intensity within 𝑅𝑂𝐼_𝑙𝑒𝑠𝑖𝑜𝑛_ , and 𝑆𝐼_𝑏𝑟𝑎𝑖𝑛_ denotes the mean signal intensity within 𝑅𝑂𝐼_𝑏𝑟𝑎𝑖𝑛_. The CNR and LBR were calculated for the pre-contrast, SC and CB images, while the CEP was calculated for the SC and CB images, with respect to the pre-contrast image. Percentage increase of CNR, LBR and CEP was calculated between SC and CB images.

### Qualitative Assessment

Three board-certified radiologists , , and , with 9, 15 and 18 years of experience respectively, were presented with the imaging sets of 110 cases each consisting of the pre-contrast, SC and CB images. The readers viewed SC and CB images side-by-side, while using the pre-contrast as an optional reference image. Reader scored both the SC and CB images using a 4-point Likert scale for each lesion visualization and image quality endpoints as shown in Table 2. Additionally, readers were also asked to score whether the blood vessel conspicuity was increased, decreased or as expected or if they impact diagnosis. Table 2 summarizes the scoring criteria for the different qualitative assessment endpoints.

**Table 2:**
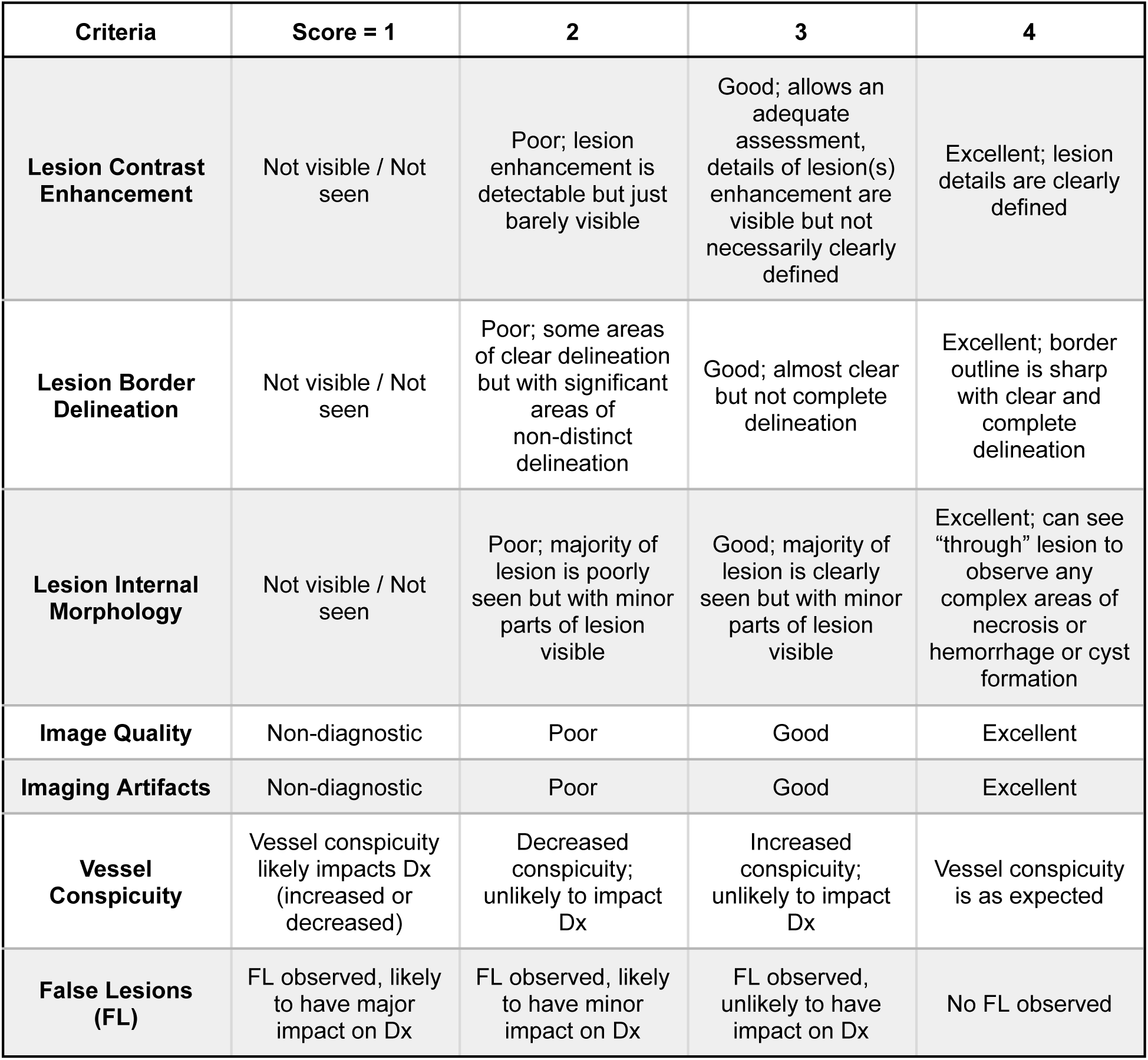
Scoring criteria for the different qualitative assessment criteria. Dx - Diagnosis.

| Criteria | Score = 1 | 2 | 3 | 4 |
| --- | --- | --- | --- | --- |
| **Lesion Contrast Enhancement** | Not visible / Not seen | Poor; lesion enhancement is detectable but just barely visible | Good; allows an adequate assessment, details of lesion(s) enhancement are visible but not necessarily clearly defined | Excellent; lesion details are clearly defined |
| **Lesion Border Delineation** | Not visible / Not seen | Poor; some areas of clear delineation but with significant areas of non-distinct delineation | Good; almost clear but not complete delineation | Excellent; border outline is sharp with clear and complete delineation |
| **Lesion Internal Morphology** | Not visible / Not seen | Poor; majority of lesion is poorly seen but with minor parts of lesion visible | Good; majority of lesion is clearly seen but with minor parts of lesion visible | Excellent; can see "through" lesion to observe any complex areas of necrosis or hemorrhage or cyst formation |
| **Image Quality** | Non-diagnostic | Poor | Good | Excellent |
| **Imaging Artifacts** | Non-diagnostic | Poor | Good | Excellent |
| **Vessel Conspicuity** | Vessel conspicuity likely impacts Dx (increased or decreased) | Decreased conspicuity; unlikely to impact Dx | Increased conspicuity; unlikely to impact Dx | Vessel conspicuity is as expected |
| **False Lesions (FL)** | FL observed, likely to have major impact on Dx | FL observed, likely to have minor impact on Dx | FL observed, unlikely to have impact on Dx | No FL observed |

### Statistical Analysis

Quantitative metrics and qualitative scores were compared for statistical significance using the Wilcoxon signed rank test. The different P-values used are mentioned along with the respective results. All statistical analysis was performed using Python v3.7 with the SciPy v1.14 library.

## Results

The quantitative and qualitative assessment was done based on the 110 patients (47 ± 22 years; 52 Females, 47 Males, 11 N/A), selected as per the inclusion criteria and the power calculation shown in Figure 1. Detailed patient demographics can be found in Table 1.

### Quantitative Assessment

CB images had better quantitative performance when compared to SC images with a significant increase in CNR (729.17%), LBR (87.91%), and CEP (165.81%). Mean and standard deviation for all quantitative endpoints are provided in Table 3.

**Table 3:** Contrast-to-noise Ratio (CNR), Lesion-to-brain Ratio (LBR) and Contrast Enhancement Percentage (CEP) shown for all cases and lesional cases. The percentage increase is calculated from SC to CB and (*) denotes that the increase in values from SC to CB is statistically significant with a p < 0.001 (Wilcoxon signed rank test).

|  | All Cases |  |  | Lesion Cases |  |  |
| --- | --- | --- | --- | --- | --- | --- |
|  | CNR | LBR | CEP | CNR | LBR | CEP |
| Pre-contrast | 4.31 ± 3.79 | 0.88 ± 0.41 | - | 4.07 ± 3.27 | 0.83 ± 0.43 | - |
| Standard Contrast (SC) | 10.25 ± 11.33 | 3.33 ± 1.65 | 213 ± 173 % | 6.13 ± 9.25 | 2.53 ± 0.96 | 108 ± 72% |
| Contrast Boost (CB) | 50.75 ± 69.90* | 5.72 ± 2.30* | 390 ± 259%* | 32.22 ± 41.49* | 4.60 ± 2.04* | 226 ± 113%* |
| Percentage Increase | 729.17 ± 1576.92% | 87.91 ± 72.12% | 165 ± 423% | 1120.20 ± 2534.10% | 80.20 ± 40.60% | 151.10 ± 108.60% |

**Table 4:**
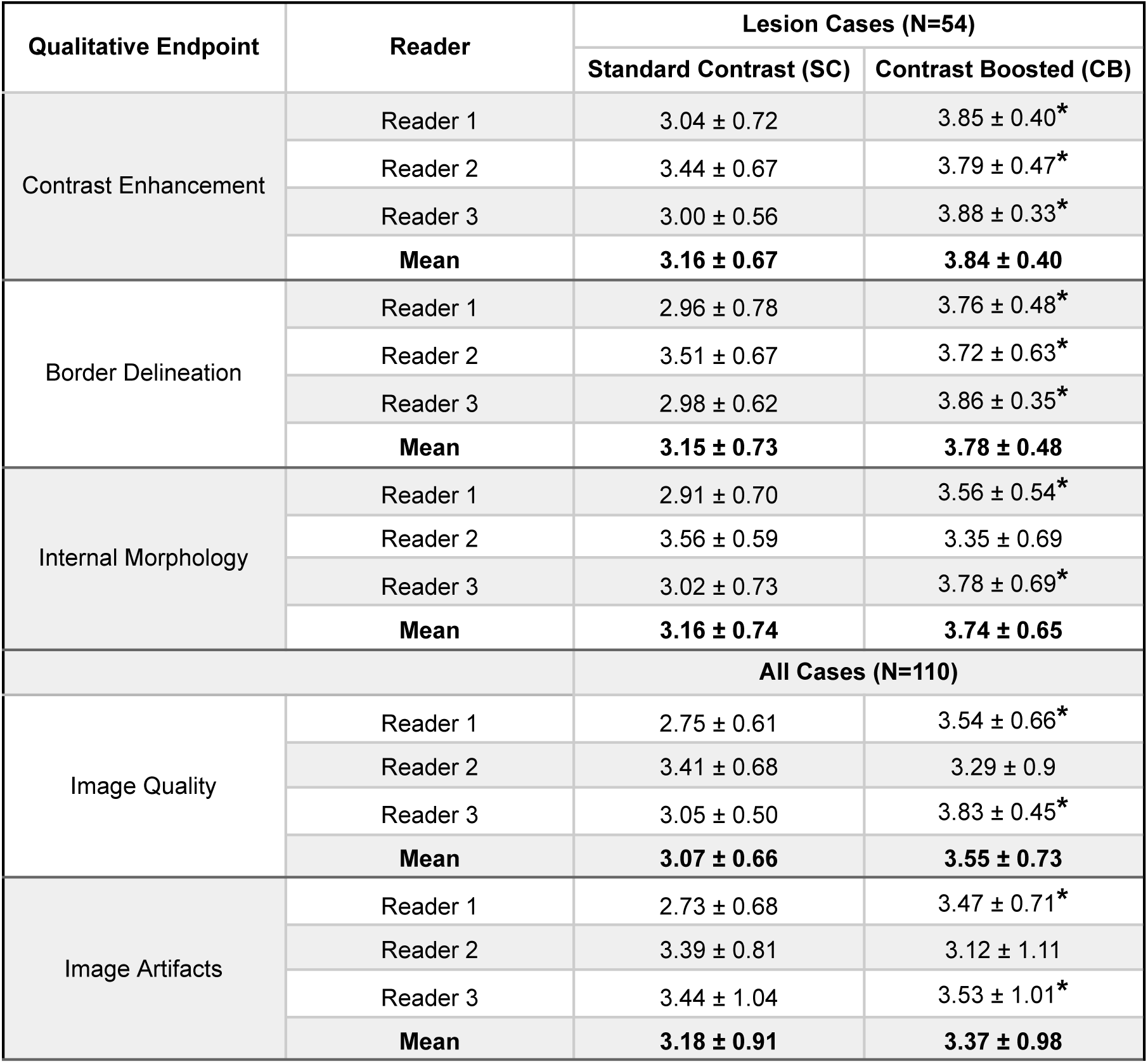
Summary of reader scores for the different qualitative endpoints described in Table 2. Scores for the lesion specific endpoints (contrast enhancement, border delineation and internal morphology) are shown only for lesional cases while image quality and artifact scores are shown for all cases. (*) denotes statistical significance with p < 0.001 (Wilcoxon signed rank test).

### Qualitative Assessment

Fig. 3 shows examples of pre-contrast, SC and CB images for different clinical indications. CB images were compared quantitatively and qualitatively with respect to SC images. For lesional cases, CB images have significantly better lesion contrast enhancement (3.84 vs 3.16), lesion border delineation (3.78 vs 3.15) and lesion internal morphology (3.56 vs 3.16) when compared to SC images. Fig. 4 visualizes the qualitative performance of CB images with respect to SC images for the different lesion visualization endpoints. CB images also have significantly better overall image quality (3.55 vs 3.07) and significantly less imaging artifacts (3.37 vs 3.18).

**Fig 3:**
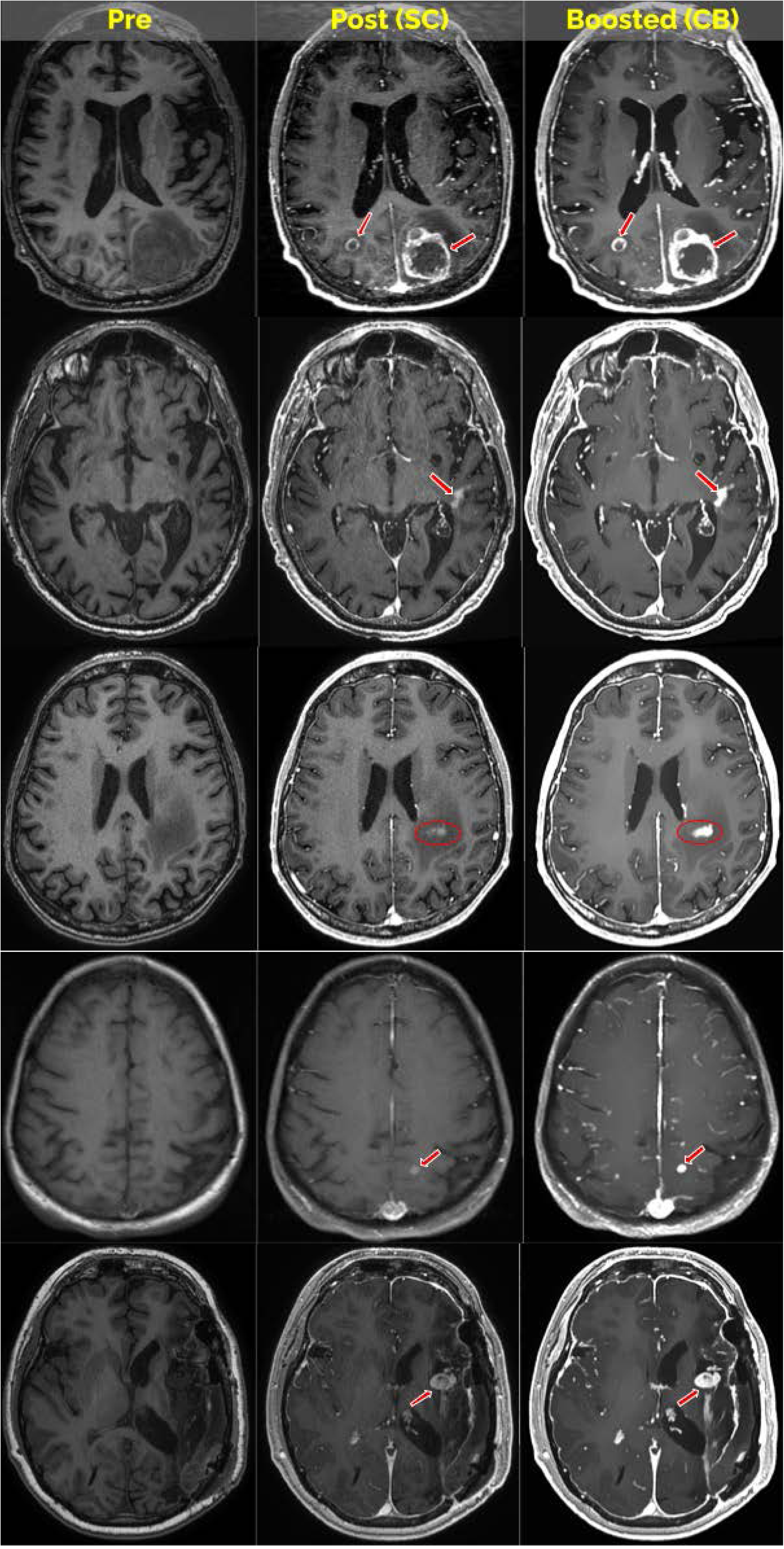
Pre-contrast, standard-contrast and contrast boosted images for five different patients with different clinical indications.

**Fig 4:**
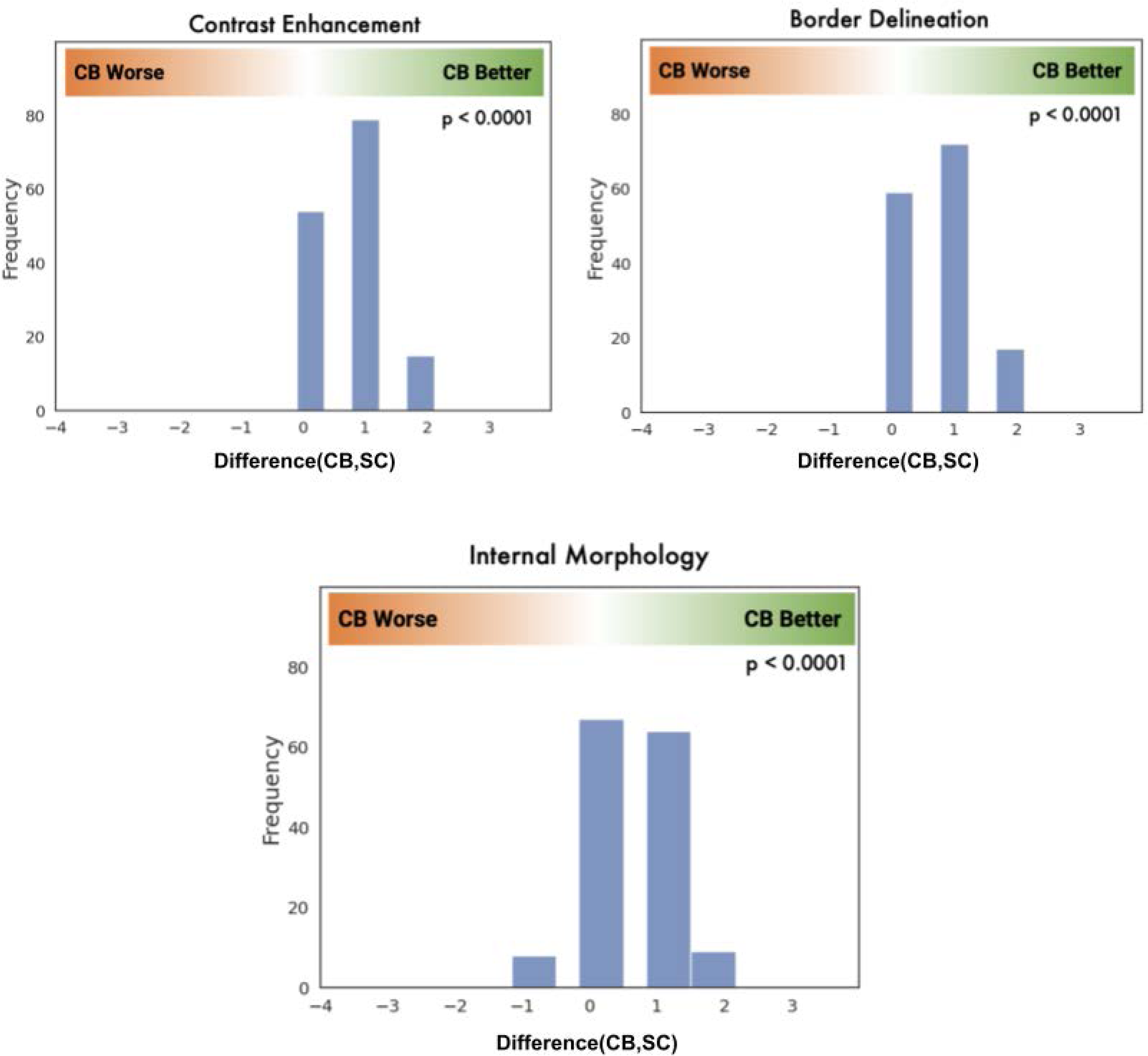
Comparison of contrast boosted images with respect to standard-contrast images on different lesion specific qualitative endpoints shown only for lesional cases for each read (330 reads in total - 110 cases times 3 readers).

Vessel Conspicuity on CB images was assessed to be likely to impact diagnosis in 20.00% of cases compared to 3.03% for SC, as shown in Fig. 5. It was observed that vessel conspicuity was a matter of reader preference with some readers finding the increased vessel conspicuity more impactful than others.

**Fig 5:**
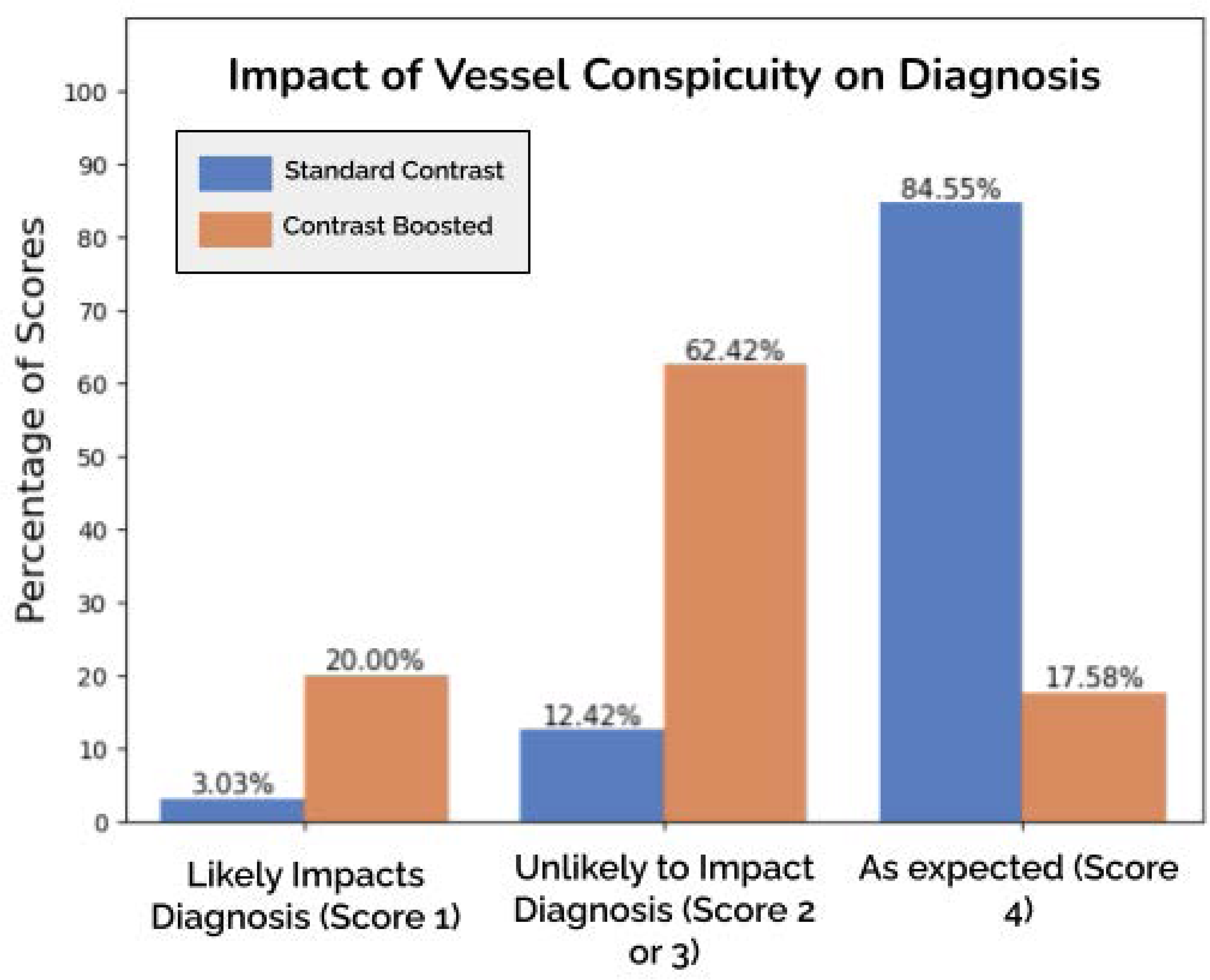
Likelihood of vessel conspicuity impacting diagnosis, averaged over the three readers.

To analyze the frequency, cause and severity of potential false lesions seen on CB images, readers were asked to note any CB lesions they suspected were false positive based on the pre-contrast and SC images. Readers were asked to assess the potential of false positive lesions for impacting diagnosis as unlikely, minor or major impact. Readers indicated if they could attribute the false positive lesions to errors or artifacts from the input images or if the false positive was introduced by the DL algorithm, or both. Finally, readers were asked whether or not they could confidently rule out the contrast-boosted lesion as a false positive using the corresponding SC image.

From the false positive lesion analysis, it was observed that CB images may introduce false positive lesions due to registration errors between the input sequences, image artifacts (i.e., motion, susceptibility, and flow artifacts), vessel conspicuity introduced by the boosting algorithm itself, or a combination of these causes. Fig. 6 summarizes the frequency and severity of false lesions in the CB images. In 100% of cases readers reported that the SC image was sufficient to confidently rule out lesions as being false positives. These results provide evidence that the potential impact of false lesions is circumscribed to a small portion of cases and can be identified as false positive using the SC image.

**Fig 6:**
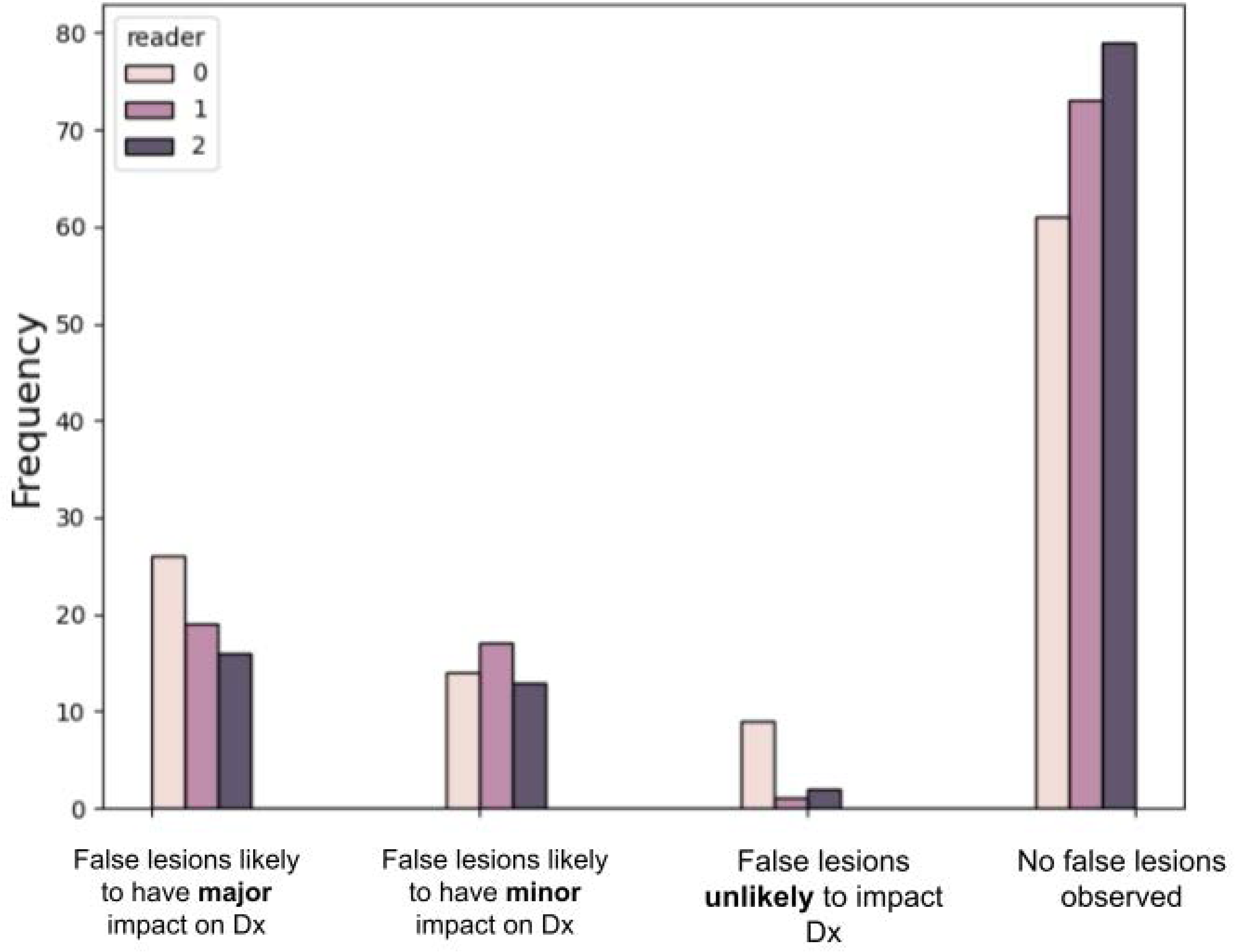
Frequency and severity of false lesions observed on contrast-boosted images. Dx - Diagnosis.

### Dose Concentration Appearance

We used the following physics-based method to calculate dose concentration on the SC and CB images, to establish that CB images have a dose concentration appearance that is equivalent to images obtained with higher than standard-of-care dosage levels (0.1 mmol/kg for all and 0.05 mmol/kg for Gadopiclenol) for a given contrast agent with relaxivity 𝑟_1_.

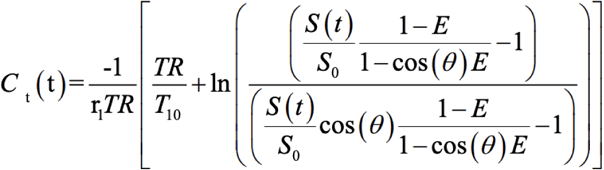

where C is dose concentration, r_1_ is the relaxivity constant of Gd chelate, TR is repetition time, T_10_ is baseline tissue T_1_ time, E = e^−TR/T10^, θ is the acquisition flip angle (FA), S is signal intensity, and S_0_ - pre-contrast signal intensity.

The physics-based dose concentration calculation showed a 92.5 ± 62.7% increase over SC images (range: 5.8-383.8%), showing that the contrast enhancement in CB images is approximately equivalent to the contrast enhancement expected after administration of a double dose of a contrast agent.

## Discussion

The results of this multi-reader multi-center retrospective study demonstrates that DL based contrast boosting yields better lesion visualization and overall image quality through quantitative and qualitative evaluation. We quantitatively established that the contrast information present in the CB images is superior to that of the SC images.

The high increase of CNR (729.17%) of CB images compared to SC images is due to the fact that both the contrast between a lesion and the normal parenchyma is increased by CB and also the noise of the images is effectively reduced by the denoising module of the algorithm. By definition, CNR is inversely proportional to the standard deviation of the signal intensity in the normal brain, which tends to vary between the individual cases. These factors contribute to the relatively high standard deviation of the CNR values. The LBR and CEP metrics are not affected by the denoising module and are not sensitive to the factors influencing the variability described above. Both LBR and CEP of CB images are significantly better (87.91% and 165.81%, respectively) than that of the SC images. Both metrics are measuring the main effects of contrast boosting.

For lesion contrast enhancement assessed in all patients with identifiable lesions, all readers demonstrated a higher preference in favor of CB (average score 4 - “Excellent”; 3.79- 3.88) over SC images (3 - “Good”; 3.00 - 3.44). Similarly for lesion border delineation all readers demonstrated an increased preference for CB (average score 4; 3.72-3.86) over SC images (average score 3; 2.98-3.51). For lesion internal morphology, two readers demonstrated an increased preference for CB images while one reader demonstrated no preference. CB images were rated between 3 and 4 with a range of 3.35 to 3.78 while SC image scores ranged from 2.91 to 3.56. For IQ, all readers showed a better average score for CB (3.3 to 3.8) images when compared to SC images (2.8 to 3.4). One reader found a higher mean score of image artifacts for the CB images, meaning a lower presence and clinical impact of artifacts, while two readers demonstrated no significant difference. Overall, CB provides a significant reduction of artifacts compared to SC. As CB images have contributions of information from both pre-contrast and SC images, any artifacts present in only one image are reduced in the final results. This along with the denoising module achieves the effect of artifact reduction.

Though vessel conspicuity is higher in most of the CB images, it does not impact diagnosis on the majority of the cases, with some variability between readers. In post-hoc conversations with readers, the difference in preference was attributed to their experience using vessel suppressed sequences at their institution. Overall, in approximately 80% of cases vessel conspicuity in CB images was unlikely to impact diagnosis. Therefore the potential negative impact of the higher vessel conspicuity with contrast boosting is limited. Future studies will be dedicated to reducing the vessel conspicuity without affecting contrast boosting.

Through an extensive analysis of false lesions, we have established that the majority of false lesions introduced in CB images do not have any impact on the diagnosis and that they can easily be ruled out using the SC images as reference. Using a physics-based method for calculating dose concentration we showed that CB images are approximately equivalent to double dose contrast enhanced images.

Our findings are consistent with the analysis performed by Bône et. al., [24]. The dose appearance analysis further builds on these results and strengthens the fact that the CB images can be used in place of double-dose images. Haase et. al., [28] recently published a deep learning based signal amplification algorithm with true double dose as reference. The percentage increase achieved in this proposed work is better than that of the artificial double dose in [28] in terms of CNR(729% vs 253%), LBR(87% vs 42%) and CEP(165% vs 120%).

This study has several limitations. The DL algorithm is sensitive to input image quality and registration. Poor registration and image artifacts can introduce false positive lesions into CB images. Although physics-guided analysis showed that CB images are equivalent to double-dose images, the availability of actual double-dose images is necessary to establish equivalent clinical performance. Future studies will evaluate CB images using acquired double-dose images as ground truth. Though we have evaluated the diagnostic performance of DL-based contrast boosting through a side-by-side evaluation, future studies will evaluate the diagnostic interchangeability through blinded reads.

In conclusion, the quantitative and qualitative results show that DL-based contrast boosting has significantly superior lesion visualization performance when compared to standard-dose contrast enhanced brain MRI images and can be used in applications that require higher lesion sensitivity, without the need to increase the administered contrast dose.

## Data Availability

All data produced in the present study are proprietary and are not available to public

---

Supplementary Fig 1: Comparison of CNR between the training and performance datasets. Similar CNR patterns show that contrast-boosted images are equivalent to higher-dose contrast-enhanced images.

## References

1. Lohrke, J., Frenzel, T., Endrikat, J., Alves, F. C., Grist, T. M., Law, M., … & Pietsch, H. (2016). 25 years of contrast-enhanced MRI: developments, current challenges and future perspectives. Advances in therapy, 33, 1–28.

2. Ellingson, B. M., Harris, R. J., Woodworth, D. C., Leu, K., Zaw, O., Mason, W. P., … & Cloughesy, T. F. (2017). Baseline pretreatment contrast enhancing tumor volume including central necrosis is a prognostic factor in recurrent glioblastoma: evidence from single and multicenter trials. Neuro-oncology, 19(1), 89–98.

3. Koudriavtseva, T., Pozzilli, C., Gasperini, C., Argentino, C., Di Biasi, C., Iannilli, M., … & Gualdi, G. F. (1996). High-dose contrast-enhanced MRI in multiple sclerosis. Neuroradiology, 38, S5–S9.

4. Montagne, A., Nation, D. A., Pa, J., Sweeney, M. D., Toga, A. W., & Zlokovic, B. V. (2016). Brain imaging of neurovascular dysfunction in Alzheimer’s disease. Acta neuropathologica, 131, 687–707.

5. Sheldon, J. J., Siddharthan, R., Tobias, J., Sheremata, W. A., Soila, K., & Viamonte, M. (1985). MR imaging of multiple sclerosis: comparison with clinical and CT examinations in 74 patients. American journal of neuroradiology, 6(5), 683–690.

6. Aljahdali, S., Azim, G., Zabani, W., Bafaraj, S., Alyami, J., & Abduljabbar, A. (2024). Effectiveness of radiology modalities in diagnosing and characterizing brain disorders. Neurosciences Journal, 29(1), 37–43.

7. Martucci, M., Russo, R., Schimperna, F., D’Apolito, G., Panfili, M., Grimaldi, A., … & Gaudino, S. (2023). Magnetic resonance imaging of primary adult brain tumors: state of the art and future perspectives. Biomedicines, 11(2), 364.

8. Fink, J. R., Muzi, M., Peck, M., & Krohn, K. A. (2015). Multimodality brain tumor imaging: MR imaging, PET, and PET/MR imaging. Journal of Nuclear Medicine, 56(10), 1554–1561.

9. Nazarpoor, M., Poureisa, M., & Daghighi, M. H. (2012). Comparison of maximum signal intensity of contrast agent on t1-weighted images using spin echo, fast spin echo and inversion recovery sequences. Iranian Journal of Radiology, 10(1), 27.

10. Subedi, K. S., Takahashi, T., Yamano, T., Saitoh, J. I., Nishimura, K., Suzuki, Y., … & Nakano, T. (2013). Usefulness of double dose contrast-enhanced magnetic resonance imaging for clear delineation of gross tumor volume in stereotactic radiotherapy treatment planning of metastatic brain tumors: a dose comparison study. Journal of radiation research, 54(1), 135–139.

11. Radbruch, A., Weberling, L. D., Kieslich, P. J., Hepp, J., Kickingereder, P., Wick, W., … & Bendszus, M. (2015). High-signal intensity in the dentate nucleus and globus pallidus on unenhanced T1-weighted images: evaluation of the macrocyclic gadolinium-based contrast agent gadobutrol. Investigative radiology, 50(12), 805–810.

12. Rogowska, J., Olkowska, E., Ratajczyk, W., & Wolska, L. (2018). Gadolinium as a new emerging contaminant of aquatic environments. Environmental toxicology and chemistry, 37(6), 1523–1534.

13. McDonald, R. J., McDonald, J., & Therneau, T. (2017). Assessment of the neurologic effects of intracranial gadolinium deposition using a large population based cohort. Illinois: Radiological Society of North America.

14. Robic, C., Port, M., Rousseaux, O., Louguet, S., Fretellier, N., Catoen, S., … & Corot, C. (2019). Physicochemical and pharmacokinetic profiles of gadopiclenol: a new macrocyclic gadolinium chelate with high T1 relaxivity. Investigative Radiology, 54(8), 475–484.

15. Loevner, L. A., Kolumban, B., Hutóczki, G., Dziadziuszko, K., Bereczki, D., Bago, A., & Pichiecchio, A. (2023). Efficacy and safety of gadopiclenol for contrast-enhanced MRI of the central nervous system: the PICTURE randomized clinical trial. Investigative radiology, 58(5), 307–313.

16. Hofmann, B. M., Riecke, K., Klein, S., Klemens, M. A., Palkowitsch, P., Kahn, J. F., … & Ebert, W. (2023). Relationship of Dose and Signal Enhancement Properties of Gadoquatrane, a New Tetrameric, Macrocyclic Gadolinium-Based Contrast Agent, Compared With Gadobutrol: A Randomized Crossover Study in Healthy Adults. Investigative Radiology, 10-1097.

17. Manning, P., Daghighi, S., Rajaratnam, M. K., Parthiban, S., Bahrami, N., Dale, A. M., … & Farid, N. (2020). Differentiation of progressive disease from pseudoprogression using 3D PCASL and DSC perfusion MRI in patients with glioblastoma. Journal of Neuro-Oncology, 147, 681–690.

18. Ross, J. S., Buckner Petty, S. A., Brinjikji, W., Hoxworth, J. M., Lehman, V. T., Middlebrooks, E. H., … & Wood, C. P. (2020). Multiple reader comparison of 2D TOF, 3D TOF, and CEMRA in screening of the carotid bifurcations: Time to reconsider routine contrast use?. Plos one, 15(9), e0237856.

19. Haase, R., Pinetz, T., Bendella, Z., Kobler, E., Paech, D., Block, W., … & Deike-Hofmann, K. (2023). Reduction of gadolinium-based contrast agents in MRI using convolutional neural networks and different input protocols: limited interchangeability of synthesized sequences with original full-dose images despite excellent quantitative performance. Investigative radiology, 58(6), 420–430.

20. Pasumarthi, S., Tamir, J. I., Christensen, S., Zaharchuk, G., Zhang, T., & Gong, E. (2021). A generic deep learning model for reduced gadolinium dose in contrast-enhanced brain MRI. Magnetic Resonance in Medicine, 86(3), 1687–1700.

21. Kleesiek, J., Morshuis, J. N., Isensee, F., Deike-Hofmann, K., Paech, D., Kickingereder, P., … & Radbruch, A. (2019). Can virtual contrast enhancement in brain MRI replace gadolinium?: a feasibility study. Investigative radiology, 54(10), 653–660.

22. Liu, J., Pasumarthi, S., Duffy, B., Gong, E., Datta, K., & Zaharchuk, G. (2023). One model to synthesize them all: Multi-contrast multi-scale transformer for missing data imputation. IEEE transactions on medical imaging, 42(9), 2577–2591.

23. Wang, D., Pasumarthi, S., Zaharchuk, G., & Chamberlain, R. (2023, October). Simulation of arbitrary level contrast dose in MRI using an iterative global transformer model. In International Conference on Medical Image Computing and Computer-Assisted Intervention (pp. 88–98). Cham: Springer Nature Switzerland.

24. Bône, A., Ammari, S., Menu, Y., Balleyguier, C., Moulton, E., Chouzenoux, É., … & Lassau, N. (2022). From dose reduction to contrast maximization: can deep learning amplify the impact of contrast media on brain magnetic resonance image quality? A reader study. Investigative Radiology, 57(8), 527–535.

25. Mingo, A. F., Serra, S. C., Macula, A., Bella, D., La Cava, F., Alì, M., … & Valbusa, G. (2023). Amplifying the effects of contrast agents on magnetic resonance images using a deep learning method trained on synthetic data. Investigative Radiology, 58(12), 853–864.

26. Kharaji, Mona, et al. "DANTE-CAIPI Accelerated Contrast-Enhanced 3D T1: Deep Learning–Based Image Quality Improvement for Vessel Wall MRI." American Journal of Neuroradiology 46.1 (2025): 49–56.

27. Lewis, D., Zhu, X., Coope, D. J., Zhao, S., King, A. T., Cootes, T., … & Li, K. L. (2022). Surrogate vascular input function measurements from the superior sagittal sinus are repeatable and provide tissue-validated kinetic parameters in brain DCE-MRI. Scientific Reports, 12(1), 8737.

28. Haase, R., Pinetz, T., Kobler, E., Bendella, Z., Zülow, S., Schievelkamp, A. H., … & Deike, K. (2024). Deep Learning–Based Signal Amplification of T1-Weighted Single-Dose Images Improves Metastasis Detection in Brain MRI. Investigative Radiology, 10-1097.

