## Supplementary for "Deep-Learning Based Contrast Boosting Improves Lesion Visualization and Image Quality: A Multi-Center Multi-Reader Study on Clinical Performance with Standard Contrast Enhanced MRI of Brain Tumors"

**Article Type:** Original Research

#### **Supplementary Material**

#### Lesion Size Measurement

To establish the size range of lesions analyzed in this study, two neuroimaging researchers (__., and __) segmented the largest lesion in pathological cases using itk-SNAP [1]. In cases where the two lesion segmentation volumes differed by less than 20%, the first reader's value was reported. If lesion sizes differed by more than 20%, a third independent adjudicator J.S., (neuroradiologist with 17 years of experience) selected the most appropriate segmentation. The range of enhancing lesions sizes were reported and a sub-group analysis of lesion contrast enhancement was performed for small (< 1cc) and large (> 1cc) lesions.

The analysis included a wide range of lesion sizes (0.02-59.8 cc), with an average lesion size of 4.2 cc (95% confidence interval 1.2-7.1 cc). Additionally, a subgroup analysis was performed to demonstrate algorithm performance on small lesions (N=17, lesion <1cc) and large lesions (N=26, lesion >1cc). For both small and large lesions, contrast enhancement ratings by readers were greater in contrast-boosted images than in standard-contrast images.

### Radiomics Feature Analysis

To analyze the impact of the algorithm on the texture content of enhancing lesions of CB images, we used PyRadiomics (version 3.1.0) to compare 120 radiomics features[2]. These features were computed on $ROI_{lesion}$ and grouped into six feature classes: first-order statistics, gray level co-occurrence matrix (GLCM), gray level run length matrix (GLRLM), gray level size zone matrix (GLSZM), neighboring gray-tone difference matrix (NGTDM), and gray level dependence matrix (GLDM). Pixel values inside the ROIs were normalized and binned using a bin width of 25, as recommended by PyRadiomics. We computed the concordance correlation coefficient (CCC) for each of the six feature classes. The CCC ($\hat{\rho}_c$) is defined as:

$$\hat{\rho}_c = \frac{2s_{xy}}{s_x^2 + s_y^2 + (\bar{x} - \bar{y})^2}$$

where $\bar{x}$ and $\bar{y}$ are the mean values, $s_x^2$ and $s_y^2$ are the variances, and $s_{xy}$ is the covariance defined as:

$$s_{xy} = \frac{1}{N}\sum_{n=1}^{N}(x_n - \bar{x})(y_n - \bar{y})$$

The correlation of six radiomics feature classes of CB and SC images are tabulated in Supplementary Table 1. All features have a good concordance correlation coefficient (CCC) (mean CCC: 0.78 ± 0.05) for lesions and excellent correlation (mean CCC: 0.88 ± 0.08) for parenchyma tissue / healthy tissue when comparing similarity in texture between SC and CB images.

| Radiomic Feature | CCC Lesion | CCC Parenchyma |
|---|---|---|
| First Order | 0.91 ± 0.02 | 0.86 ± 0.02 |
| Gray Level Co-occurrence Matrix (GLCM) | 0.68 ± 0.03 | 0.84 + 0.03 |
| Gray Level Dependence Matrix (GLDM) | 0.73 ± 0.06 | 0.92 + 0.10 |
| Gray Level Run Length Matrix (GLRLM) | 0.76 ± 0.08 | 0.89 ± 0.11 |
| Gray Level Size Zone Matrix (GLSZM) | 0.78 ± 0.07 | 0.88 ± 0.09 |
| Neighbouring Gray Tone Difference Matrix (NGTDM) | 0.81 ± 0.02 | 0.91 ± 0.12 |
| **Mean** | **0.78 ± 0.04** | **0.88 ± 0.08** |

Supplementary Table 1: Mean and standard deviations of concordance correlation coefficient (CCC) for the six radiomic features shown for lesion and parenchymal regions. Lesion radiomic features have a good concordance with CCC=0.78 whereas parenchymal regions have an excellent concordance with CCC=0.88.

### CNR Analysis

The change in CNR between pre-contrast, low dose post-contrast, and SC images in the training set was compared to the change in CNR between pre-contrast, SC, and CB images in the evaluation dataset. For the training dataset, similar ROIs are drawn and the CNR of pre-contrast, low-dose and full-dose images are calculated and plotted.

Supplementary Figure 1. illustrates the CNR analysis between the training and evaluation datasets. The CNR increase follows a similar pattern between the two datasets, with a similar slope, showing that CB images are equivalent to higher-dose CE images. In other words, the CNR increase in the training set for a dose level increase from 0.01 mmol/kg (low-dose) to 0.1 mmol/kg (SC) is equivalent to the CNR increase in the performance set for a dose level increase from 0.1 mmol/kg (SC) to the dose level in CB images. Hence it can be concluded that the dose level in CB images is much higher than that of SC images.

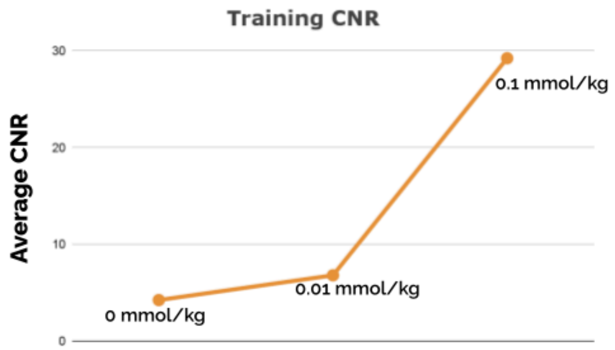


Supplementary Fig 1: Comparison of CNR between the training and performance datasets. Similar CNR patterns show that contrast-boosted images are equivalent to higher-dose contrast-enhanced images.

### References

1. Yushkevich, P. A., Piven, J., Hazlett, H. C., Smith, R. G., Ho, S., Gee, J. C., & Gerig, G. (2006). User-guided 3D active contour segmentation of anatomical structures: significantly improved efficiency and reliability. Neuroimage, 31(3), 1116-1128.
2. Van Griethuysen, J. J., Fedorov, A., Parmar, C., Hosny, A., Aucoin, N., Narayan, V., ... & Aerts, H. J. (2017). Computational radiomics system to decode the radiographic phenotype. Cancer research, 77(21), e104-e107.
